# Investigating the contribution of circulating inflammatory cytokines on the link between obesity and COVID-19

**DOI:** 10.1101/2025.02.07.25321874

**Authors:** Zahra J. Khamis, Emmanouil Karteris, Amani Alhajeri, Steven G. Smith, Alexandra Blakemore, Fotios Drenos

**Affiliations:** Department of Life Sciences, College of Health, Medicine and Life Sciences, Brunel University London, London, UK; Government Hospitals, The Genetics Center, Manama, Kingdom of Bahrain; Department of Metabolism, Digestion and Reproduction, Imperial College London, London, UK; College of Medicine, Nursing, and Health Science, University of Galway, Galway, Republic of Ireland

**Keywords:** COVID-19, SARS-CoV2, inflammatory cytokines, mediation analysis, Two-sample Mendelian randomization (2SMR), Multivariable Mendelian randomization (MVMR), Janus kinase 2 (JAK2), leptin, lymphocytes percentage

## Abstract

**Background:** Coronavirus disease 2019 (COVID-19), is more severe in individuals with obesity. Furthermore, a cytokine storm was observed in many critically ill patients with COVID-19. Since adipose tissue secretes cytokines, we investigated whether cytokines mediate the effect of obesity on COVID-19 severity.

**Methods:** Using two-sample Mendelian randomization analyses we assessed the causal effect of body mass index (BMI) on COVID-19 severity. We then evaluated the BMI effect on 41 inflammatory cytokines, as well as on JAK-2, lymphocyte percentage and leptin. We also tested the relationship between these immunological factors and COVID-19 severity. The estimates obtained were used in a mediation analysis to understand the immunological factors linking BMI to COVID-19 severity.

**Results:** Higher BMI increased the risk of COVID-19 severity. BMI was also causally associated with 5 of the 41 inflammatory cytokines, HGF, TRAIL, IL 13, IL6, and IL 7. We identified TNF-α and IL-8 as the only two inflammatory cytokines associated with COVID-19 severity. Leptin-related genetic variation was associated with COVID-19 severity, but JAK-2 and lymphocyte percentage were not. We found no statistical evidence of mediation of immunological factors tested on the relationship between BMI and COVID-19 severity, although our estimate was 52.8%.

**Conclusions:** We replicated the previously reported association between BMI and COVID-19 severity. We identified the inflammatory cytokines elevated due to higher BMI. Other inflammatory cytokines showed evidence for increasing COVID-19 severity. However, we were unable to find statistical evidence of baseline levels of circulating cytokines, or additional factors involved in a cytokine storm i.e. JAK-2, lymphocyte percentage, and leptin, mediating the link between BMI and severe COVID-19. Although targeting specific cytokines will be of benefit in the general population, further work on cytokine levels during early phases of COVID-19 infection is needed to provide new approaches to decrease the risk of severe COVID-19 with higher BMI.

## 1 Background

Obesity, the excessive accumulation of fat, is considered to be one of the major risk factors for developing severe COVID-19 (1). Several studies indicate that, as body mass index (BMI) increases, so does the risk of mechanical ventilation and ICU admission (2)

Adipose tissue (AT), where body fat is stored, is an active immune organ that regulates hormonal, metabolic, and immune processes. AT plays an important role in the development of systemic and local low-grade inflammation that is recognized by local infiltration of immune cells, and by elevated circulating proinflammatory factors (3).

According to our current understanding, following infection with SARS-CoV2, resident macrophages are activated in adipose tissue. M1-polarized macrophage recruitment promotes the production of local and systematic pro-inflammatory cytokines (IL 8, IL 6, TNF α) resulting in a type-1 immune response (Th1) (4). Individuals with obesity develop a chronic immune response in adipose tissue, resulting in the production of B cells, T cells, and natural killer (NK) cells that can produce a cytokine storm. A cytokine storm is the result of the activity of several immune cells including dendritic cells, macrophages, natural killer cells and T and B lymphocytes. Upon viral infection, the innate immune cells recognize the molecular structure of the invading virus, called pathogen associated molecular patterns (PAMPs), by pattern recognition receptors (PRRs) around the cells. The interaction between PRRs and PAMPs triggers signaling pathways that induce the expression of genes encoding proinflammatory cytokines (5). The sudden increase in cytokines, "the cytokine storm", results in accumulation of immune cells into the infection site, causing tissue damage. This can include damage to endothelial cells, the vascular barrier, and its capillaries, as well as diffuse alveolar damage, which may lead to multiorgan failure, and death (6).

The immune response cascade is also affected by other key players such as: leptin; Janus kinase 2 (JAK2); and lymphocytes (7). As well as regulating appetite and reproductive functions, leptin is a proinflammatory adipokine secreted by adipose tissue. Thus, leptin contributes to inflammation and may lead to chronic inflammation (7). It directly stimulates macrophages (M1 phenotype), natural killer (NK) cells, dendritic cells (DC) and T-helper cells to secrete proinflammatory cytokines such as IL1, IL6, and TNFα (8).

The Janus kinase (JAK)/signal transducer and activator of transcription (STAT) pathway is the most important signaling pathway of the leptin receptor. Upon leptin binding, JAK2 activation occurs, which leads to STAT3 binding to phosphorylated tyrosine in leptin receptor followed by expression of responsive genes (9). JAK2 might be imbalanced in patients with obesity could lead to an exaggerated immune response (9).

Lymphocytes, mainly CD4+ T cells and CD8+ T-cells ; play an essential role as antiviral immunity (10). Patients who had recovered from COVID-19 had high to normal levels of T-cells (11). In severe cases of SARS-CoV2, patients have low levels of lymphocytes, mainly CD8+ (10). Therefore, lymphocyte counts have been suggested as possible biomarkers to evaluate the severity of COVID-19 in patients (11).

The link between obesity and COVID-19 severity has been hypothesized as being, at least partly, due to the immune responses of visceral fat in patients with obesity (6). There have been several reports describing the effects of inflammatory cytokines on COVID-19 infection (12) and the role of obesity in increasing the severity of COVID-19 (10,11).

To test the mediating role of cytokines and related immunological markers in the relationship between BMI and COVD-19 severity, we need to first understand the causal relevance and direction of effects between the different factors. Mendelian randomization (MR) allows us to use genetic variants associated with an exposure of interest as instrumental variables to assess causality. When used as a two-sample approach, summary results from large genome-wide association studies (GWAS) can be used, significantly increasing the number of people we can include in the analysis and, thus, improving our statistical power to detect a modest effect on an outcome (13).

In this study, using publicly-available data from exceptionally large GWAS including hundreds of thousands of individuals and the Mendelian randomization methodology, we aim to investigate the potential mediation role of 41 cytokines, leptin, JAK2, and lymphocyte percentage in the link between obesity and COVID-19 severity. Identifying the mediators of the effect of obesity on COVID-19 severity will allow us to provide epidemiological evidence for targeting specific cytokines that might decrease the risk of COVID-19 on patients with obesity, and which might also have wider consequences for other categories of patients.

## 2 Materials and Methods

### 2.1. Study Design

The study was performed through three MR analyses as outlined in Figure (1): (1) MR analysis was used to estimate the causal association between BMI and severe COVID-19. (2) We also used MR analysis to test the effect of BMI on circulating cytokines and related traits. (3) We conducted MR analysis to identify the circulating cytokines and related traits causally relevant to COVID-19 severity. Finally, we tested whether any of the factors identified as associated with BMI and COVID-19 severity shows evidence of a mediating role between the two, using a multivariable MR and estimating the possible mediating effect.

### 2.2. Selection of genetic instrument variables for BMI

The genetic instruments for BMI were extracted from the Metabochip meta-analysis and genome-wide association study conducted by Locke, *et al*. The study identified 289 BMI-associated loci (at a p-value threshold of < 5 × 10 ^-8^) from 322,154 participants of European descent (14).

### 2.3. GWAS summary statistics for COVID-19 severity

COVID-19 severity genetic association data were obtained from the COVID-19 host genetics initiative (COVID-19 HG) GWAS meta-analysis (15) (https://www.COVID-19hg.org/). The COVID-19 HG team has made summary data publicly available on multiple COVID-19 outcomes from several studies. The data acquired for this analysis is with an accession ID: “GCST011073” at the GWAS Catalog of European populations. The study is from May 13, 2020, which is the last time point available before the wide release of the COVID-19 vaccination. Severity of infection is defined as hospitalized COVID-19 Vs. not hospitalized COVID-19 release 5 and includes 38,984 European ancestry cases and 1,644,784 European ancestry controls. The relevant data can be downloaded from (https://www.ebi.ac.uk/gwas/studies/GCST011073 ) (15).

### 2.4. GWAS summary statistics for inflammatory cytokines and C-reactive protein

The summary statistics for inflammatory cytokines were acquired from the work of Kalaoja, *et al.* (12). The authors analyzed a cytokine panel (41 cytokines) from the National FINRISK Study (1997 and 2002 cohorts) and the Cardiovascular Risk in Young Finns Study (YFS) The full GWAS summary results for all 41 inflammatory cytokines are available through the University of Bristol (https://data.bris.ac.uk/data/dataset/3g3i5smgghp0s2uvm1doflkx9x) (16).

### 2.5. GWAS summary statistics for leptin levels, lymphocytes percentage, and Tyrosine-protein kinase JAK2 levels

The summary statistics for lymphocyte percentage of leukocytes were obtained from the study by Jung H, et. al. (17) based on data of 174,488 individuals from the UK biobank with European ancestry (U.K.). Lymphocyte percentage was measured by dividing the number of lymphocytes in blood by the total number of white blood cells.

Summary statistics data for leptin levels were acquired from the published study by Yaghootkar H. *et al.* (18) accession ID: “GCST90007310” and downloaded from the GWAS Catalog https://www.ebi.ac.uk/gwas/studies/GCST90007310. Yaghootkar analyzed circulating leptin levels in 49,909 European ancestry individuals for association with exonic genetic variants (18).

GWAS of tyrosine-protein kinase (JAK2) levels was obtained from a genome-wide association study of serum proteins published by Gudjonsson. et.al (19), accession ID: “GCST90086866” and was downloaded from the GWAS Catalog https://www.ebi.ac.uk/gwas/studies/GCST90086866. The authors performed a GWAS for 5,355 European Icelandic ancestry individuals with 2,091 serum proteins including JAK2 measured through the SOMAmer platform (19).

### 2.6. Statistical Analysis

All analyses were performed using R (20). The TwoSampleMR package (21) was used for MR and the ggplot2 package (22) for plots, unless otherwise stated. The inverse variance weighted (IVW) approach was considered as the main MR analysis for estimating the causal relationships unless there was evidence for directional pleiotropy. Directional pleiotropy was tested through the intercept term of the MR Egger analysis method (23). In the presence of pleiotropy, we used the weighted median estimate. All p-values with P <0.05 were considered statistically significant. All association estimates were reported as either odd ratios (OR) with 95% confidence intervals (CIs) for binary outcomes, or as beta coefficients and 95% CIs for continuous outcome variables. The mediation analysis was performed as described in the supplementary material of Burgess et al (24). Causal Mediation analysis is a method that assesses a hypothetical causal effect of exposure and outcome via a mediator. It separates the total effect into direct and indirect effect whereby the indirect effect includes the hypothetical mediator to the outcome. The total effect is obtained from the two main sample MR results. The direct effect of BMI on COVID-19 severity was estimated using a Multivariable Mendelian randomization using the MVMR package in R. Multivariable MR analysis allows the simultaneous estimation of causal effects of multiple exposures on an outcome (25). The indirect effect can then be estimated as the difference between the total and direct effects *(Figure 4)*.

## 3 Results

### 3.1 The causal effect of BMI on COVID-19 severity

From the previously reported 289 BMI associated SNPs, 187 remained after removing variants with linkage disequilibrium of r^2^ > 0.001 with any other SNP. All 187 were found in the COVID-19 severity data and were successfully harmonized between the two sets of results. The inverse variance weighted (IVW) estimates showed that BMI was positively associated with COVID-19 hospitalization (OR=1.242; 95% CI 1.034 -1.317, p=1.63 x10^-8^). MR Egger was used to test the existence of pleiotropy, and no evidence of directional pleiotropy was present (Intercept 0.0005, *p*-value = 0.638).

### 3.2 The effect of BMI on inflammatory cytokines

We estimated the effect of BMI on 41 inflammatory cytokines. Genetically-predicted BMI was positively associated with five inflammatory cytokines with P-value less than 0.05 as shown in Supplementary Table 1 and Figure 2. The cytokine with the highest magnitude of effect from BMI was hepatocyte growth factor (HGF) (β = 0.277, se = 0.0703, *p*=8.00 x 10^-5^). Other cytokines affected included TNF-related apoptosis-inducing ligand (TRAIL) (β = 0.221, se = 0.0678, *p=*1.09 x10^-3^), IL 13 (β = 0.226, se = 0.0916, *p=*1.38 x10^-2^), IL 6 (β = 0.148, se = 0.0638, *p=*2.01 x10^-2^) and IL 7 (β = 0.196, se = 0.0992, *p=*4.83 x10^-2^). None of the cytokines tested showed evidence of decreasing with increasing BMI as shown in Figure 2 and Supplementary Table 1, though CTACK was close (b = -0.184, se =0.097, p = 5.6×10^-2^).

**Figure 1.**
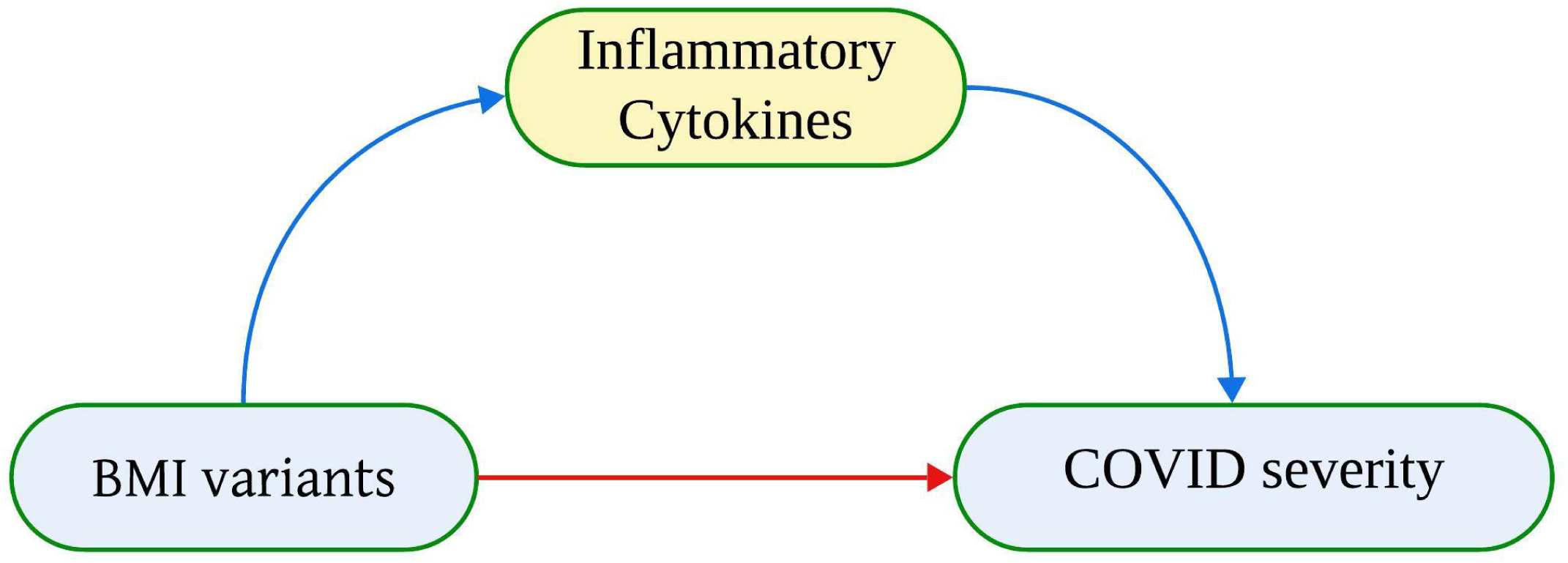
A simplified illustration of total effect, direct, and indirect effect using two sample Mendelian randomization. Direct effect between exposure (BMI variants) and outcome (COVID-19 severity) is C’. Indirect effect between exposure and outcome through mediator (cytokines) is a’+b’. The total effect is Direct effect + Indirect effect.

**Figure 2.**
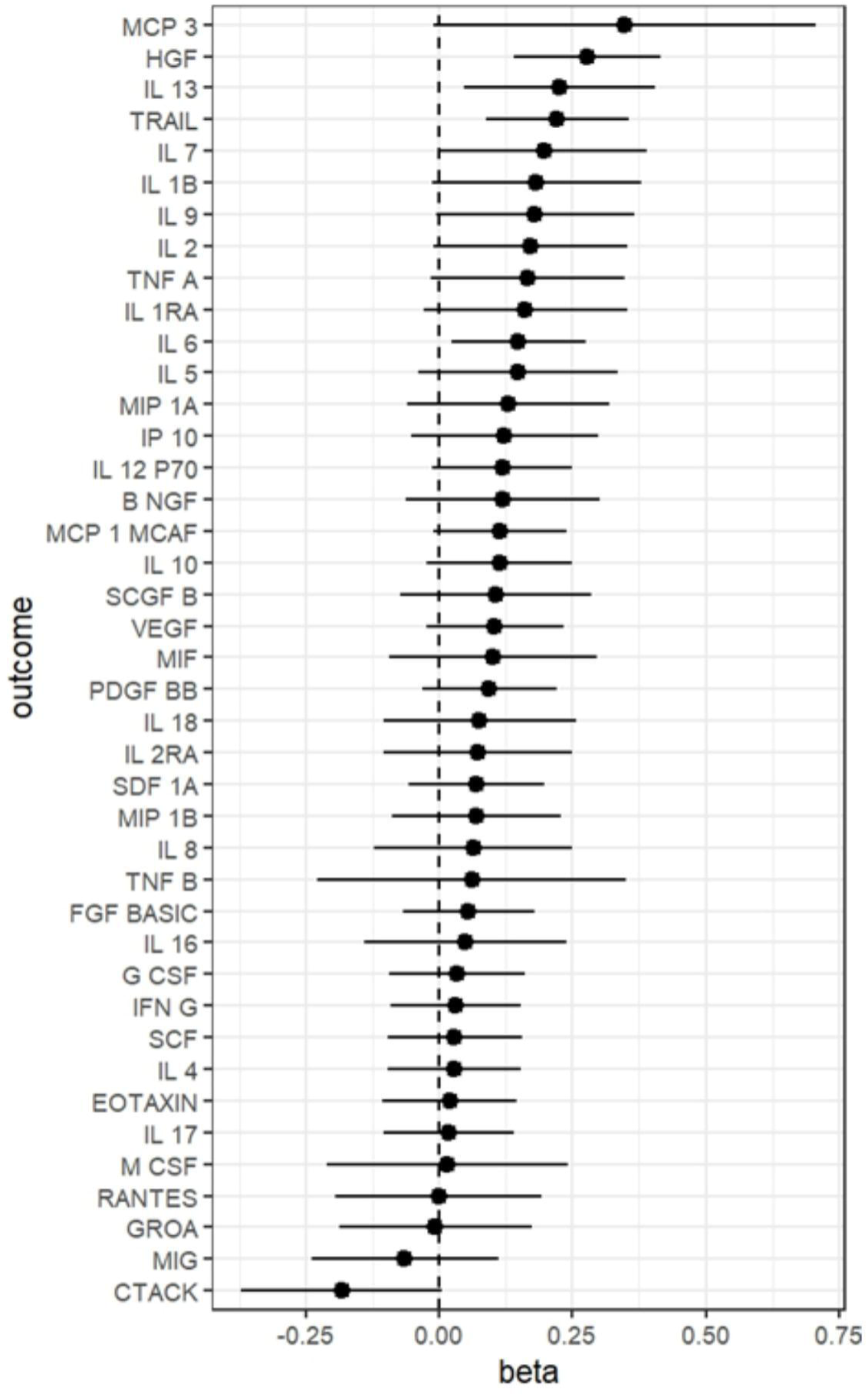
·Causal effect (betas) from the Two Sample Mendelian randomization analysis of body mass index (BMI) (exposure) with infl.ammatory cytokines (outcome)

### 3.3 The effect of inflammatory cytokines on COVID-19 severity

We tested all 41 inflammatory cytokines for a causal association with COVID-19 severity. Of these, two cytokines were positively associated with severity of COVID-19: TNF α (OR= 1.030 (95% CI 1.011-1.048, *p*=1.72 x 10^-3)^ and IL 8 (OR=1.017; 95% CI 1.000 - 1.034, *p=*4.88 x10^-2)^. On the other hand, HGF (OR=0.892; 95% CI 0.800-0.984, *p=*1.46 x10^-2^) was the only cytokine with evidence of being inversely associated with COVID-19 severity as shown in Figure 3 and Supplementary Table 2. We found no evidence of directional pleiotropy for any of the associations tested using the intercept of the MR Egger model (Figure 3 and Supplementary Table 2).

**Figure 3.**
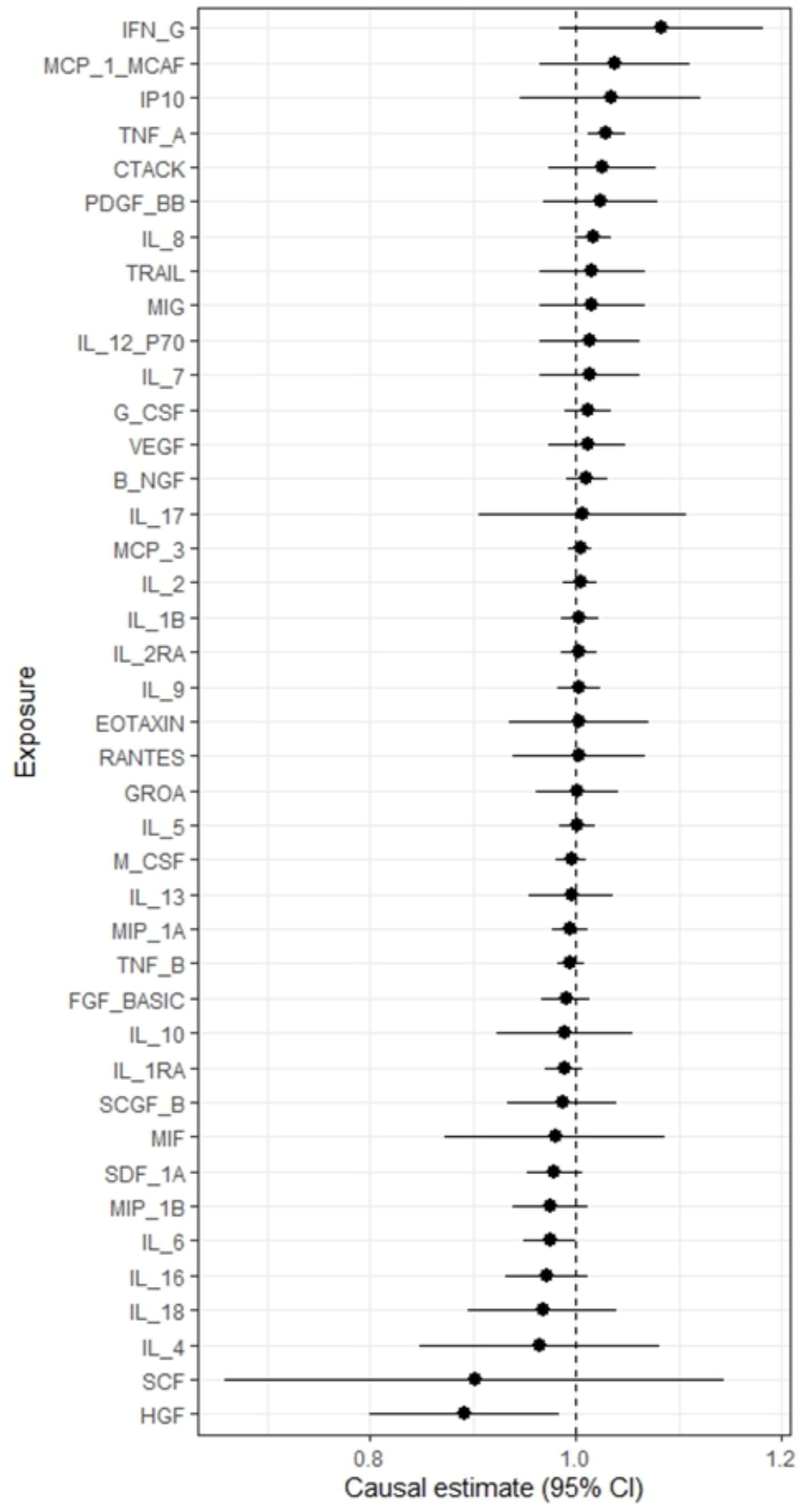
Causal effect estimates (odds ratios and 95% confidence intervals) from the Two-Sample Mendelian randomization analysis of inflammatory cytokins (exposure) with COVID-19 severity (outcome).

### 3.4 The effect of BMI on each of leptin, JAK2, and lymphocyte percentage

We analyzed the effect of BMI variants individually on leptin, JAK2, and lymphocyte percentage. Genetically-predicted BMI was only statistically significantly associated with leptin (β = 0.732, se = 0.0798, *p=*4.756 x10^-20^). JAK2 (β = 0.142, se = 0.101, *p=*0.16) and lymphocytes (β = 0.0475, se = 0.0308, *p=0.*12) showed no evidence of associations.

### 3.5 The effect of leptin, JAK2, and lymphocyte percentage on COVID-19 severity

Leptin-associated genetic variants were positively associated with COVID-19 severity (OR=1.289; 95% CI: 1.0766-1.501, *p=*1.92 x10^-2^), whilst no statistically significant evidence was found between SNPs for JAK2 (OR= 1.006 (95% CI 0.950 – 1.061, *p*=0.83) and lymphocyte percentage (OR= 0.989 (95% CI 0.908-1.069, *p*=0.78) with COVID-19 severity.

### 3.6 Mediation Analysis

Since no single cytokine measure was associated with both BMI and COVID-19 severity. We measured the indirect causal effect of BMI on COVID-19 severity via all inflammatory cytokines together. Leptin was associated with both BMI and COVID-19 severity, and we also tested its mediation effect *(Figure 4)*.

**Figure 4.**
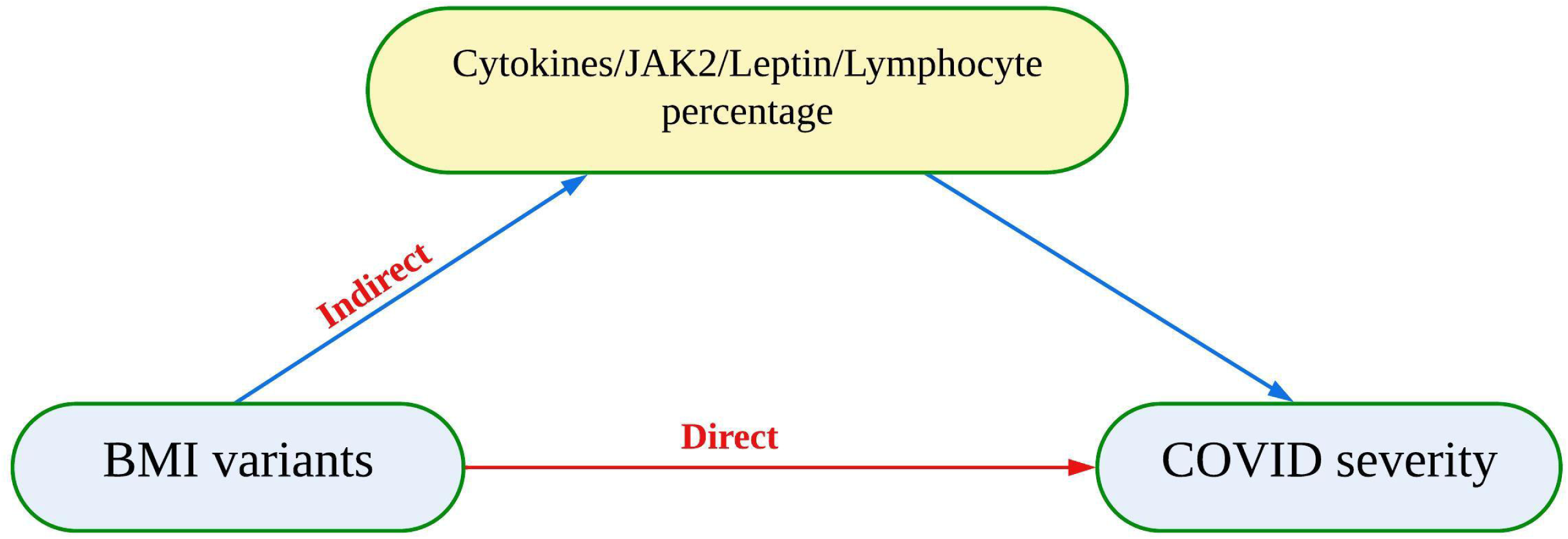
A simplified illustration of mediation analysis, direct, and indirect effect using two sample mendelian randomization. Direct effect between exposure (BMI variants) and outcome (COVID-19 severity). Indirect effect between exposure and outcome through mediator (cytokines, JAK2, Leptin, and lymphocytes percentage).

#### 3.6.1 MVMR of BMI and all inflammatory cytokines on COVID-19 severity

To examine the possible mediation effect of all inflammatory cytokines together, and due to the strong correlation between cytokines, we followed a stepwise approach in building our MVMR model by adding inflammatory cytokines, guided by the knowledge of immune response pathways to identify those contributing independent information and exclude those causing collinearity. Four non-correlated cytokines were included in the final model, BNGF, GROA, IL2RA, and MIP1A*(Supplementary Table 3)*. The direct effect of BMI (103 SNPs) on COVID-19 severity was no longer statistically significant (OR= 0.969 (95% CI 0.812 – 1.125) (*Supplementary Table 3*, but none of the inflammatory cytokines was statistically significant with COVID-19 severity (*Supplementary Table 3)*.

#### 3.6.2 MVMR of BMI and leptin levels on COVID-19 severity

When leptin was included as a conditional factor in the BMI to COVID-19 severity MR model through MVMR, the direct effect of BMI (26 SNPs) on COVID-19 severity was no longer statistically significant (OR= 0.9839 (95% CI 0.752 – 1.216).

### 3.7 Causal Mediation Analysis

The mediation of the joint inflammatory cytokines was tested. The BMI effect mediated by the cytokines included in the model was estimated as 0.102 (CI -0.0542, 0.2585), but the confidence intervals include 0, indicating lack of statistical significance for the estimate (*Supplementary Table 4)*. This represents 52.8% of the total BMI effect estimated, but despite its large value we were not able to find statistical evidence of a mediation effect. Mediation analysis was performed for leptin as it showed evidence of positive association with both BMI and COVID-19. We re-estimated the total effect using the BMI SNPs from the MVMR and leptin causal mediation analysis also showed no evidence of a significant association, indirect effect of 0.129 (CI - 0.1592, 0.4172). Again, although this was 19.7% of the BMI effect on COVID-19 severity, we lack the statistical evidence to confirm its presence (*Supplementary Table 3*).

## 4 Discussion

Several studies have identified obesity, particularly as measured by high body mass index (BMI), as a risk factor for COVID-19 severity. Since obesity also promotes inflammation, the contribution of cytokines in increasing the risk of severe COVID-19 has been hypothesized, but is not yet well understood.

Severe forms of COVID-19 are also associated with an overreaction of the immune system through a cytokine storm. Here, using a two sample Mendelian randomization approach we confirmed the causal link between obesity, as measured by high BMI, with COVID-19 severity in the form of hospitalization following an infection. We also established the causal effects of genetically-predicted BMI on cytokines and of cytokines to COVID-19 severity. We investigated the effect of BMI on leptin, JAK2 and lymphocyte percentage and their effect on COVID-19 severity. Mediation analysis was performed for the examined risk factors on the link between BMI and COVID-19 severity and although a large proportion of the link between the two can potentially be explained by a combination of cytokines and leptin, we were not able to find clear statistical evidence to support their mediating role.

Our results agree with both observational and MR studies done previously between BMI and COVID-19 severity: persons with obesity are at higher risk of severe course pf COVID infection. The effect we estimated (OR = 1.207; 95% CI: 1.115–1.307; ) is close to that reported by several Mendelian randomization studies of BMI and COVID-19 severity. COVID-19 severity in high BMI patients was also observed in a previously published meta-analysis of 167 studies including 3 million COVID-19 patients. The meta-analysis was performed to compare the association between high BMI hospitalized patients and normal BMI COVID-19 hospitalized patients and the outcome results showed that severity of COVID-19 is 1.5 times higher in obese patients compared to non-obese patients. The slight overestimation of the effect of BMI indicates the possible presence of confounding factors between the two. Further support for the causal relationship of obesity to COVID-19 severity was seen in studies showing a decrease in COVID-19 severity in patients following bariatric surgery (26).

We assessed 41 inflammatory cytokines to investigate possible mediators that may drive a more severe reaction to the SARs-COV-2 in individuals with obesity. We found five inflammatory cytokines affected by elevated BMI levels (HGF, TRAIL, IL 13, IL 6, and IL 7). Supporting our finding, the link between obesity and changes in the cytokine profile has also been previously reported (27). Further, Dam et. al. reviewed an increase of proinflammatory cytokines; IL 1, IL 6, and TNF α by inducing immune cell infiltration from adipose tissue in cell culture settings using human primary adipocytes and found adipose cells are capable of promoting an inflammatory state (28). A two sample Mendelian randomization performed by Kalaoja et. al. identified HGF, MCP-1, and TRAIL as significantly associated with BMI variation (12).The increase of the affected cytokines seen in our study is probably due to larger number of independently significant BMI variants used compared to other studies.

We carried out two sample Mendelian randomization of inflammatory cytokines with COVID-19 severity and our results indicated causal associations with TNF α and IL 8. Both inflammatory cytokines were investigated in several studies reporting the production of high levels of several inflammatory cytokines as part of initiation of cytokine storm in COVID-19 infection such as IL 2, IL 6, IL 7, IL 8, IL 10, GSCF, IP10, MCP 1, MIP1 A, and TNF α (29–31).

We also carried out mediation analysis to investigate the role of inflammatory cytokines on the link between BMI and COVID-19 severity. We found no statistical evidence of mediation by inflammatory cytokines despite the potentially very large estimated proportion of the mediated effect. Although several studies suggested a role for inflammatory cytokines in promoting COVID-19 severity (32), and explained the role of inflammatory cytokines in obesity (27,33,34), or even explored the network effect of adipokines and cytokines in SARS-CoV-2 (35), to our knowledge, there is no published evidence of a mediating role of inflammatory cytokines on the relationship between BMI and COVID-19 severity.

Other researchers have investigated the role of leptin on obesity (36,37) and leptin in COVID-19 patients (38,39). Our study confirmed the statistically significant effect of high BMI levels on leptin and added to the evidence linking it to worsening COVID-19. Despite its known pro-inflammatory effects, we were again unable to find any statistical evidence of a mediating role of leptin in the relationship between BMI and COVID-19 severity and to our knowledge no published study has reported this effect.

We also tested JAK-2 and lymphocyte percentage, both as causal consequence of BMI and as causes of COVID-19 severity. We found no evidence for either. Sun, *et.al*. studied the causal role of different white blood cells including lymphocytes percentage and found no significant role of lymphocytes in COVID-19 severity (40). To our knowledge, no MR study has examined the role of JAK-2 with COVID-19 severity. However, JAK-2 was observed to mediate the signaling pathway of inflammatory cytokines (i.e.: IL 6)(41) and anti-JAK-2 inhibitors are suggested for use to mitigate SARs-CoV-2 infection (42,43).

MR, since it uses genetic variants that commonly explain only a small part of the variation of the exposure of interest, frequently require very large sample sizes to identify the modest associations present (44) which has likely affected our ability to obtain statistical evidence for mediation despite the high proportion of the effect explained. As GWAS efforts increase in size and more genetic variants are identified, we may be able to confirm this effect in the future. This will require GWAS for cytokines with comparable size to the ones available for BMI and COVID-19, though the difficulty and cost in obtaining these measures may make this challenging. Our work contains additional limitations. A major issue for all MR studies is pleiotropy. To ensure the validity of our results, we used the MR-Egger method to test for the presence of directional pleiotropy. The results indicated that our findings are unaffected by it, but pleiotropy remains a concern in all MR studies, more so when positive effects are reported. In two-sample MR approaches, it is important to use information from comparable populations as systematic changes between them may influence the results (45). Here, our analysis is necessarily restricted to people of European ancestry, due to relevant GWAS data availability. It is unclear whether our conclusions hold for other ancestries, as some studies reported differences in the genetic architecture of the traits considered in this work among different ancestries, particularly in inflammatory cytokines secretion and function. Studying healthy individuals in GWAS might also not reflect the cytokine storm initiated in viral infection. GWAS, though powerful, has limitations for studying cytokine response to viruses as it captures genetic tendency, a snapshot, rather than the dynamic response during an infection. Finally, the participants of the population study were studied in general without specification to sex. Studies showed that inflammatory cytokines and COVID-19 severity are affected by hormonal status especially in females during menopause (46) and pregnancy (47).

In our study we used two-sample Mendelian randomization to study the mediating role of inflammatory cytokines and related traits in the link between COVID-19 severity and BMI. We investigated the implications of obesity for inflammatory cytokine levels and related traits, and their effect on COVID-19 severity. We found no secure evidence of proinflammatory cytokines, or related traits, mediating the relationship between BMI and increased risk of severe COVID-19 in European populations, despite estimating a potentially large mediating effect. Given the increasing levels of obesity worldwide and the establishment of COVID-19 as an endemic infection, the effectiveness of targeting cytokines to break the link between obesity and COVID-19 severity, remains an open question. Well-powered randomized control trials will be required to confirm any suitable cytokine related interventions to control the impact of obesity on COVID-19 as done previously with the WHO Solidarity study (48). Our results in this respect remain inconclusive. Further work towards better understanding the genetics of the cytokine response, especially during early stages of infections, and the genetic mechanisms of cytokine storm initiation will help us identify potential targets for intervention not only for response to COVID-19 but also for other similar infections and potentially future pandemic-inducing infections.

## Data Availability

All data produced are available online at

https://www.COVID-19hg.org/

https://data.bris.ac.uk/data/dataset/3g3i5smgghp0s2uvm1doflkx9x

## Abbreviations

BMI: Body mass index
2SMR: Two-sample Mendelian randomization
MVMR: Multivariable Mendelian randomization
JAK2: Janus Kinase 2
IVW: inverse variance weighted approach

## Declarations

### Ethics approval and consent to participate

The work described has used published summary level data and no ethics approval is required or consent for publication.

### Consent for publication

Not applicable.

### Availability of data and materials

The data used is openly available.

### Clinical trial number

Not applicable

### Authors’ contributions

Z.K. performed the research, data acquisition, data interpretation/analysis, manuscript drafting, writing, and editing along with designing figures and creating and organizing tables. F.D. Research Concept and design and supervision, A.B. Conceptualization. F.D., A.B., E.K., A.A, and S.S. review and editing. All authors have read and approved the final manuscript.

## Acknowledgments

Not applicable

