## Supplementary tables for "Investigating the contribution of circulating inflammatory cytokines on the link between obesity and COVID-19"

**6	Supplementary data**

**Supplementary Table 1** Two sample Mendelian randomization analysis causal effect estimates of BMI (exposure) and inflammatory cytokines (outcome) using the inverse variance weighted analysis method. β:beta effect, SE: standard error~~.~~

| Exposure (BMI) | Outcome (cytokine) | | | β | SE | P-value | Egger intercept | Intercept p-value |
| --- | --- | --- | --- | --- | --- | --- | --- | --- |
| BMI | | **HGF** | 0.277 | | 0.070 | 8.00E-05 | -0.001 | 0.769 |
|  |  | **TRAIL** | 0.221 | | 0.068 | 1.09E-03 | -0.004 | 0.304 |
|  |  | **IL 13** | 0.226 | | 0.092 | 1.38E-02 | -0.003 | 0.548 |
|  |  | **IL 6** | 0.148 | | 0.064 | 2.01E-02 | 0.001 | 0.832 |
|  |  | **IL 7** | 0.196 | | 0.099 | 4.83E-02 | 0.001 | 0.816 |
|  |  | **CTACK** | -0.184 | | 0.097 | 5.60E-02 | -0.003 | 0.586 |
|  |  | **MCP 3** | 0.348 | | 0.183 | 5.73E-02 | -0.011 | 0.254 |
|  |  | **IL 9** | 0.180 | | 0.095 | 5.90E-02 | 0.002 | 0.710 |
|  |  | **IL 2** | 0.171 | | 0.093 | 6.44E-02 | -0.002 | 0.728 |
|  |  | **IL 1B** | 0.182 | | 0.100 | 6.86E-02 | 0.004 | 0.457 |
|  |  | **TNF A** | 0.166 | | 0.093 | 7.48E-02 | 0.001 | 0.842 |
|  |  | **IL 12 P70** | 0.118 | | 0.067 | 7.74E-02 | 0.002 | 0.654 |
|  |  | **MCP 1 MCAF** | 0.113 | | 0.064 | 7.79E-02 | -0.007 | 0.063 |
|  |  | **IL 1RA** | 0.162 | | 0.097 | 9.63E-02 | -0.002 | 0.684 |
|  |  | **IL 10** | 0.113 | | 0.069 | 1.04E-01 | -0.002 | 0.665 |
|  |  | **VEGF** | 0.104 | | 0.066 | 1.13E-01 | -0.005 | 0.120 |
|  |  | **IL 5** | 0.147 | | 0.096 | 1.24E-01 | -0.007 | 0.190 |
|  |  | **PDGF BB** | 0.094 | | 0.064 | 1.43E-01 | 0.000 | 0.902 |
|  |  | **IP 10** | 0.122 | | 0.090 | 1.74E-01 | -0.009 | 0.046 |
|  |  | **MIP 1A** | 0.129 | | 0.097 | 1.82E-01 | 0.003 | 0.539 |
|  |  | **B NGF** | 0.118 | | 0.093 | 2.04E-01 | -0.004 | 0.414 |
|  |  | **SCGF B** | 0.105 | | 0.091 | 2.49E-01 | 0.002 | 0.606 |
|  |  | **SDF 1A** | 0.070 | | 0.065 | 2.88E-01 | 0.002 | 0.525 |
|  |  | **MIF** | 0.100 | | 0.100 | 3.18E-01 | 0.005 | 0.331 |
|  |  | **FGF BASIC** | 0.054 | | 0.063 | 3.87E-01 | -0.002 | 0.499 |
|  |  | **MIP 1B** | 0.069 | | 0.081 | 3.98E-01 | -0.004 | 0.353 |
|  |  | **IL 18** | 0.075 | | 0.092 | 4.11E-01 | -0.007 | 0.124 |
|  |  | **IL 2RA** | 0.071 | | 0.090 | 4.30E-01 | -0.006 | 0.228 |
|  |  | **MIG** | -0.065 | | 0.090 | 4.67E-01 | -0.009 | 0.059 |
|  |  | **IL 8** | 0.064 | | 0.095 | 5.03E-01 | -0.010 | 0.048 |
|  |  | **G CSF** | 0.033 | | 0.065 | 6.11E-01 | 0.002 | 0.655 |
|  |  | **IL 16** | 0.048 | | 0.097 | 6.22E-01 | 0.003 | 0.635 |
|  |  | **IFN G** | 0.030 | | 0.063 | 6.31E-01 | -0.004 | 0.192 |
|  |  | **SCF** | 0.028 | | 0.064 | 6.58E-01 | -0.002 | 0.514 |
|  |  | **IL 4** | 0.028 | | 0.064 | 6.68E-01 | 0.000 | 0.993 |
|  |  | **TNF B** | 0.061 | | 0.148 | 6.80E-01 | 0.004 | 0.639 |
|  |  | **EOTAXIN** | 0.019 | | 0.064 | 7.68E-01 | 0.000 | 0.992 |
|  |  | **IL 17** | 0.017 | | 0.062 | 7.85E-01 | 0.000 | 0.899 |
|  |  | **M CSF** | 0.015 | | 0.116 | 8.95E-01 | 0.000 | 0.951 |
|  |  | **GROA** | -0.008 | | 0.092 | 9.34E-01 | -0.007 | 0.121 |
|  |  | **RANTES** | -0.002 | | 0.099 | 9.84E-01 | -0.004 | 0.443 |

**Supplementary Table 2** Two sample Mendelian randomization analysis causal effect estimates of inflammatory cytokines (exposure) to COVID-19 severity (outcome) using inverse variance weighted analysis method.

| Exposure  (Cytokines) | Outcome | OR | L95 | U95 | P-value | Egger intercept | Intercept p-value |
| --- | --- | --- | --- | --- | --- | --- | --- |
| TNF A | **COVID-19**  **severity** | 1.030 | 1.011 | 1.048 | 1.723E-03 | 0.578 | 0.456 |
| IL 8 |  | 1.017 | 1.000 | 1.034 | 4.876E-02 | 0.129 | 0.267 |
| IFN G |  | 1.084 | 0.985 | 1.183 | 1.113E-01 | 0.232 | 0.294 |
| SDF 1A |  | 0.979 | 0.952 | 1.007 | 1.325E-01 | 0.075 | 0.053 |
| IL 16 |  | 0.972 | 0.932 | 1.012 | 1.643E-01 | 0.424 | 0.631 |
| MIP 1B |  | 0.975 | 0.938 | 1.012 | 1.825E-01 | 0.340 | 0.278 |
| IL 1RA |  | 0.989 | 0.971 | 1.006 | 2.002E-01 | 0.456 | 0.262 |
| G CSF |  | 1.013 | 0.990 | 1.035 | 2.730E-01 | 0.202 | 0.599 |
| B NGF |  | 1.011 | 0.991 | 1.031 | 2.891E-01 | 0.896 | 0.077 |
| MCP 1 MCAF |  | 1.038 | 0.965 | 1.111 | 3.167E-01 | 0.413 | 0.216 |
| CTACK |  | 1.025 | 0.973 | 1.078 | 3.480E-01 | 0.425 | 0.446 |
| IL 18 |  | 0.968 | 0.895 | 1.040 | 3.770E-01 | 0.766 | 0.118 |
| PDGF BB |  | 1.024 | 0.968 | 1.080 | 4.061E-01 | 0.904 | 0.954 |
| FGF BASIC |  | 0.990 | 0.967 | 1.014 | 4.150E-01 | 0.665 | 0.728 |
| MCP 3 |  | 1.005 | 0.994 | 1.015 | 4.158E-01 | 0.317 | 0.562 |
| TNF B |  | 0.995 | 0.982 | 1.008 | 4.433E-01 | 0.720 | 0.680 |
| IP 10 |  | 1.034 | 0.947 | 1.121 | 4.546E-01 | 0.636 | 0.440 |
| VEGF |  | 1.012 | 0.974 | 1.049 | 5.410E-01 | 0.273 | 0.355 |
| TRAIL |  | 1.016 | 0.965 | 1.068 | 5.420E-01 | 0.118 | 0.038 |
| IL 4 |  | 0.965 | 0.849 | 1.081 | 5.492E-01 | 0.134 | 0.348 |
| M CSF |  | 0.996 | 0.981 | 1.010 | 5.606E-01 | 0.808 | 0.278 |
| MIP 1A |  | 0.995 | 0.977 | 1.012 | 5.667E-01 | 0.395 | 0.168 |
| IL 12 P70 |  | 1.014 | 0.965 | 1.062 | 5.804E-01 | 0.984 | 0.821 |
| IL 2 |  | 1.004 | 0.988 | 1.021 | 6.043E-01 | 0.832 | 0.483 |
| SCGF B |  | 0.987 | 0.934 | 1.040 | 6.289E-01 | 0.789 | 0.389 |
| IL 1B |  | 1.004 | 0.986 | 1.022 | 6.659E-01 | 0.938 | 0.079 |
| MIF |  | 0.980 | 0.874 | 1.087 | 7.154E-01 | 0.579 | 0.154 |
| IL 9 |  | 1.004 | 0.983 | 1.024 | 7.344E-01 | 0.276 | 0.010 |
| EOTAXIN |  | 1.003 | 0.936 | 1.071 | 9.219E-01 | 0.719 | 0.855 |
| GROA |  | 1.002 | 0.961 | 1.042 | 9.337E-01 | 0.833 | 0.303 |
| HGF |  | 0.892 | 0.800 | 0.984 | 1.458E-02 | 0.564 | 0.530 |
| MIG |  | 1.016 | 0.964 | 1.067 | 5.505E-01 | 0.301 | 0.345 |
| SCF |  | 0.902 | 0.659 | 1.145 | 4.041E-01 | 0.456 | 0.000 |
| IL 2RA |  | 1.004 | 0.987 | 1.020 | 6.849E-01 | 0.734 | 0.300 |
| IL 5 |  | 1.001 | 0.984 | 1.018 | 8.781E-01 | 0.737 | 0.315 |
| IL 6 |  | 0.975 | 0.950 | 1.000 | 4.523E-02 | 0.688 | 0.852 |
| IL 7 |  | 1.014 | 0.965 | 1.062 | 5.836E-01 | 0.729 | 0.885 |
| IL 10 |  | 0.989 | 0.923 | 1.055 | 7.416E-01 | 0.513 | 0.317 |
| IL 13 |  | 0.996 | 0.955 | 1.037 | 8.374E-01 | 0.607 | 0.968 |
| IL 17 |  | 1.007 | 0.906 | 1.107 | 8.952E-01 | 0.683 | 0.676 |
| RANTES |  | 1.003 | 0.939 | 1.067 | 9.216E-01 | 0.443 | 0.536 |

**Supplementary Table 3** Multivariable Mendelian randomization (MVMR) of BMI and inflammatory cytokines (exposures) with COVID-19 severity (outcome). SNPs: number of single nucleotide polymorphisms; OR odds ratio; L95, U95: conﬁdence interval. BNGF: B-lymphocyte-derived neurotrophic growth factor, MIP 1A: Macrophage Inflammatory Protein 1 Alpha, GROA: Growth-Regulated Oncogene Alpha, IL 2R2: Interleukin-2 Receptor Subunit Beta.

| Exposure (BMI+ cytokines) | Outcome  (COVID-19 severity) | | OR | L95 | | U95 | | P-value | |
| --- | --- | --- | --- | --- | --- | --- | --- | --- | --- |
| BMI | | **COVID-19 (RELEASE 5)**  **ebi-a-GCST011073** | 0.968533 | | 0.812004 | | 1.125062 | | 6.89E-01 |
| BNGF | |  | 1.025432 | | 0.985036 | | 1.065828 | | 2.23E-01 |
| MIP 1A | |  | 0.972166 | | 0.920722 | | 1.02361 | | 2.82E-01 |
| GROA | |  | 1.018828 | | 0.987188 | | 1.050468 | | 2.48E-01 |
| IL 2RA | |  | 1.022909 | | 0.987461 | | 1.058356 | | 2.10E-01 |

**Supplementary Table 4** :Causal Mediation Analysis of BMI mediation analysis with noncorrelated inflammatory cytokines. β (beta) causal effect – SE: standard error, CI-: lower confidence interval, CI+: higher confidence interval. Total effect estimated of BMI SNPs after MVMR. Direct effect: effect of BMI on COVID-19 with cytokines mediation. Indirect effect: total effect-direct effect.

|  | Effect % | β | SE | CI- | CI+ |
| --- | --- | --- | --- | --- | --- |
| Total effect | 100 | 0.196143 | 0.059616 | 0.079296 | 0.31299 |
| Direct effect | 47.92% | 0.094 | 0.053 | -0.00988 | 0.19788 |
| Indirect effect | 52.08% | 0.102143 | 0.079769 | -0.0542 | 0.258489 |
